# Socio-geographic factors associated with Lyme disease in children

**DOI:** 10.64898/2026.05.15.26353361

**Authors:** Cara Wychgram, Alexandra T. Geanacopoulos, Alison W. Rebman, Laura L. Chapman, Rebecca S. Green, Desiree N. Neville, Amy D. Thompson, Meagan M. Ladell, Anupam B. Kharbanda, Kenneth D. Mandl, Frank C. Curriero, John N. Aucott, Lise E. Nigrovic, Pedi Lyme Net

**Author notes:** Corresponding author: (CW).

## Abstract

**Objective:** Lyme disease diagnosis in children is challenging due to atypical presentations and testing limitations. We sought to evaluate the association between Lyme disease and socio-geographic risk factors in children.

**Materials and methods:** We enrolled children undergoing evaluation for acute Lyme disease at one of eight Pedi Lyme Net pediatric emergency departments located in high Lyme disease incidence states over a ten-year period (2015-2024). We defined a case of Lyme disease with an erythema migrans (EM) lesion or a positive two-tier serology result in a child with signs and/or symptoms of acute disease. We linked each child’s primary residential county to the following factors: urban-rural residence, socioeconomic status, population-level disease incidence, wildland-urban interface, and “Lyme disease” Google searches. We performed a multi-level logistic regression analysis to evaluate associations between Lyme disease and county factors after adjusting for individual demographics.

**Results:** Among 5,529 children enrolled, 1,396 (25.2%) had Lyme disease: 101 (7.2%) with early-localized disease, 584 (41.8%) with early-disseminated disease, and 711 (50.9%) with late-disseminated disease. Rural residence (aOR 1.9, 95% CI 1.3–2.9), higher socioeconomic advantage (aOR 1.3, 95% CI 1.1–1.4), more “Lyme disease” Google searches (aOR 1.1, 95% CI 1.0–1.2), and higher wildland urban interface (aOR 1.2, 95% CI: 1.0–1.4) were independently associated with Lyme disease.

**Conclusion:** Incorporating socio-geographic factors alongside clinical data may augment diagnostic risk assessment in children with suspected Lyme disease. However, these factors should be incorporated carefully to ensure clinical assessments are not based on a child’s geographic location alone.

## Introduction

Lyme disease is the most common vector-borne disease in the U.S., with an estimated half a million new cases diagnosed annually [1,2]. Diagnosis requires either an erythema migrans (EM) lesion or a compatible clinical presentation confirmed by a positive two-tiered Lyme serology [3]. However, atypical clinical presentations in children and imperfect diagnostics provide challenges to accurate and timely diagnosis. Children may lack classic skin findings, and lesions may be misclassified, particularly in children with darker skin [4–7]. Serologies may be falsely negative early in disease and often require days to result, resulting in diagnostic delays [8].

As Lyme disease is spatially heterogeneous, contextual geographic information that refines clinician pretest probability may augment initial decision-making while awaiting diagnostic test results [9]. Area-level measures of rurality, tick habitat presence, socioeconomic status, and population-level disease incidence have been previously associated with Lyme disease risk [9–19].**Error! Bookmark not defined.Error! Bookmark not defined.** Recent work using Google data has linked geographic patterns in internet searches to disease incidence and expansion [20,21]. To date, these associations remain underexplored both at the patient-level and in children, who are at high risk for Lyme disease [22].

Understanding how socio-geographic factors influence pediatric Lyme disease risk may support future development of clinical decision support tools. However, practically speaking, candidate factors should be easily accessible to clinicians, who lack geospatial expertise. To this end, we selected readily obtainable socio-geographic factors and evaluated their associations with Lyme disease in a multi-center prospective cohort of children with suspected acute Lyme disease.

## Materials and methods

### Study design, setting, and population

We assembled a prospective cohort of children 1 to 21 years of age who presented to one of eight Pedi Lyme Net emergency departments located in high Lyme disease incidence states over a ten-year period (2015–2024) [23]. Study staff approached eligible children with either a clinician-diagnosed EM lesion or Lyme disease serology ordered for study participation. Details of the study protocol were described previously [23]. We obtained written informed consent from caregivers with assent as appropriate and consent if > 18 and <= 21 years of age. Enrollment started on June 3, 2015 and ended on October 31, 2024. The Institutional Review Boards of each participating center approved the study with permission for data sharing.

### Patient and visit characteristics

We approached caregivers of (< 18 years) and enrolled patients (age 18 to 21 years) to collect demographics and providers for clinical history and physical exam findings at the time of enrollment. Using National Institutes of Health guidance [24], we included self-reported race (American Indian/Alaskan Native, Asian, Black, Pacific Islander, White, and Other) and ethnicity (Hispanic or non-Hispanic) due to racial and ethnic differences in Lyme disease incidence [5–7]. To account for small groups, we analyzed race and ethnicity in the following four categories: White non-Hispanic, Black non-Hispanic, Hispanic and Other. Based on our observations at the enrolling centers, we defined peak Lyme disease season as presentation between June and October [25]. We then abstracted diagnostics tests, treatment, and clinical outcome form the medical record and by phone call one month after enrollment.

### Primary outcome

Our primary outcome was a diagnosis of Lyme disease defined by either a clinician-diagnosed EM lesion or a positive two-tier Lyme disease serology obtained within 30 days of enrollment in a patient with signs and/or symptoms of acute disease. Disease stage was categorized as early-localized (i.e., single EM), early-disseminated (e.g., multiple EM, facial palsy, carditis, meningitis), or late-disseminated (i.e., arthritis). All other enrolled patients with negative Lyme disease serology were classified as clinical mimics.

### Socio-geographic risk factors

Residential ZIP Code at the time of the index encounter was mapped to county based on visit date and the HUD-USPS ZIP Code Crosswalk [26]. We linked each patient’s residential county to several datasets as described in the following sections. For covariates measured at multiple time points (i.e., Social Vulnerability Index, Lyme disease incidence, and Google Trends), linkage was based on county and visit date. Covariate selection balanced our review of the literature with the desire for a concise set of variables that were easily obtainable. Broadly, we sought to capture both environmental and socio-behavioral determinants of disease risk.

### Urban-rural residence

We classified counties as urban, suburban, or rural according to the National Center for Health Statistics Urban-Rural Classification Scheme for Counties [27]. We collapsed the six-level scheme into three categories, urban (large central metro and fringe metro), suburban (medium and small metro), and rural (micropolitan and noncore) due to small group sizes among our largely urban patient population.

### Socioeconomic status

We used the socioeconomic status theme of the Social Vulnerability Index, which is calculated biennially by U.S. Centers for Disease Control and Prevention (CDC) using county measures of poverty, unemployment, housing cost burden, education, and health insurance and reported as a 0 to 1 percentile rank from least to most vulnerable counties [28]. Given previously reported associations between higher socioeconomic status and Lyme disease [10–13], we reversed the index such that higher values indicated more socioeconomic advantage.

### Lyme disease incidence

We calculated annual county incidence of Lyme disease using case counts from CDC and population estimates from the U.S. Census Bureau [29,30]. We also interactively map disease incidence on the Lyme and Tickborne Diseases Dashboard [31, 32]. When comparing incidence rates over a ten-year period, several issues arose. Given Massachusetts discontinued clinical follow-up on positive laboratory reports in 2016, resulting in very few reported cases between 2016 and 2019 [29,33], we carried forward a 2013–2015 average incidence for the missing years. Given incomplete reporting in 2020 and 2021 due to the COVID-19 pandemic [34], we carried forward the most recent complete year. We also carried 2023 data forward as CDC had not published 2024 data at the time of analysis. Lastly, after the 2022 change to laboratory-only reporting in high-incidence states [34],**^Error! Bookmark not defined.^** case counts increased sharply. To facilitate comparison across years, we normalized incidence rates by converting them to annual percentiles (0 to 100), thus ranking each county relative to all other U.S. counties in a given year.

### “Lyme disease” Google searches

We extracted annual internet search incidence for the query “Lyme disease” from the Google Trends application programming interface (API), a resource available to researchers [35]. Comparable data are mapped and downloadable from the public version of Google Trends [36], which rescales incidence rates on a range of 0 to 100 to simplify comparison across geographic locations. Incidence was expressed as the number of searches per 10 million searches by market area, which is a group of counties. We used a previously applied market area-to-county crosswalk to assign patients values [21].

### Wildland-urban interface

Wildland-urban interface indicated areas where tick habitats and human activity intersect. We obtained county measures from the Silvis Lab at the University of Wisconsin-Madison. Defined as the percentage of county area where houses meet or intermingle with wildland vegetation, wildland-urban interface identifies where human-environmental conflicts and risks are concentrated, including wildfire, habitat loss and fragmentation, invasive species, and zoonotic diseases [37–39].

### Statistical analysis

We visualized the geographic distribution of our outcome by mapping county percentages of patients with Lyme disease. We compared cases to clinical mimics using descriptive statistics.

Our primary analysis was a multi-level logistic regression with Lyme disease (vs. clinical mimic) as the outcome. In this modeling approach, individual and county fixed effects were estimated while accounting for within-county correlation via a random effect [40]. Research suggests that ignoring clustered data (e.g., children nested within counties) may lead to underestimated standard errors and biased inference [41]. We fit unadjusted models to assess each predictor individually and an adjusted model to assess independent effects. Prior to modeling, continuous variables were centered and scaled to a mean of 0 and standard deviation of 1 to make effect sizes comparable. We used a variance inflation factor < 5 to indicate no substantial multicollinearity among predictors [42]. All models included a county random effect to separate the variance in the odds of Lyme disease attributable to patient and county differences [43]. The intraclass correlation coefficient was used to assess how similar outcomes of children within the same county were, and thus whether a multi-level analysis was appropriate. A higher coefficient meant that a higher share of the total variance in the odds of Lyme disease was due to county differences. A coefficient > 0.05 (i.e., 5% of total variance) justified inclusion of a county random effect and county predictors to explain the between-county variation in Lyme disease [42–44].

Statistical analyses were performed in R version 4.5.1 and maps were produced in ArcGIS Pro version 3.1.0 [45,46].

### Sensitivity analysis

We performed a sensitivity analysis excluding population-level incidence and Google Trends from the multi-level model, as they were less analysis-ready compared to the other covariates. Due to incomplete data and case definition changes, and with no standard way to address these issues, our modifications to the population-level incidence variable were not ideal. The Google Trends data required API access, and while comparable data are publicly available on the Google Trends website, county data are not available through either data source and must be determined from larger market areas. The area-level variables in this analysis were urban-rural residence, socioeconomic status, and wildland-urban interface, all of which were publicly available by county.

## Results

From 2015 to 2024, we enrolled 5,529 children with suspected Lyme disease, of which the median age was 8 years (interquartile range: 5 to 13) and 3,131 (56.6%) were male. Overall, 1,396 (25.2%) had Lyme disease. The distribution by disease stage was as follows: 101 (7.2%) early-localized, 584 (41.8%) early-disseminated, and 711 (50.9%) late-disseminated.

Our cohort included children who resided in 200 different residential counties, with most children residing in urban counties near the enrolling study center. Among counties with at least five enrolled children, the percentage of children with Lyme disease ranged from 0 to 89% (Fig 1). In several counties in western Pennsylvania, more than 40% of enrolled children had Lyme disease.

**Fig 1.**
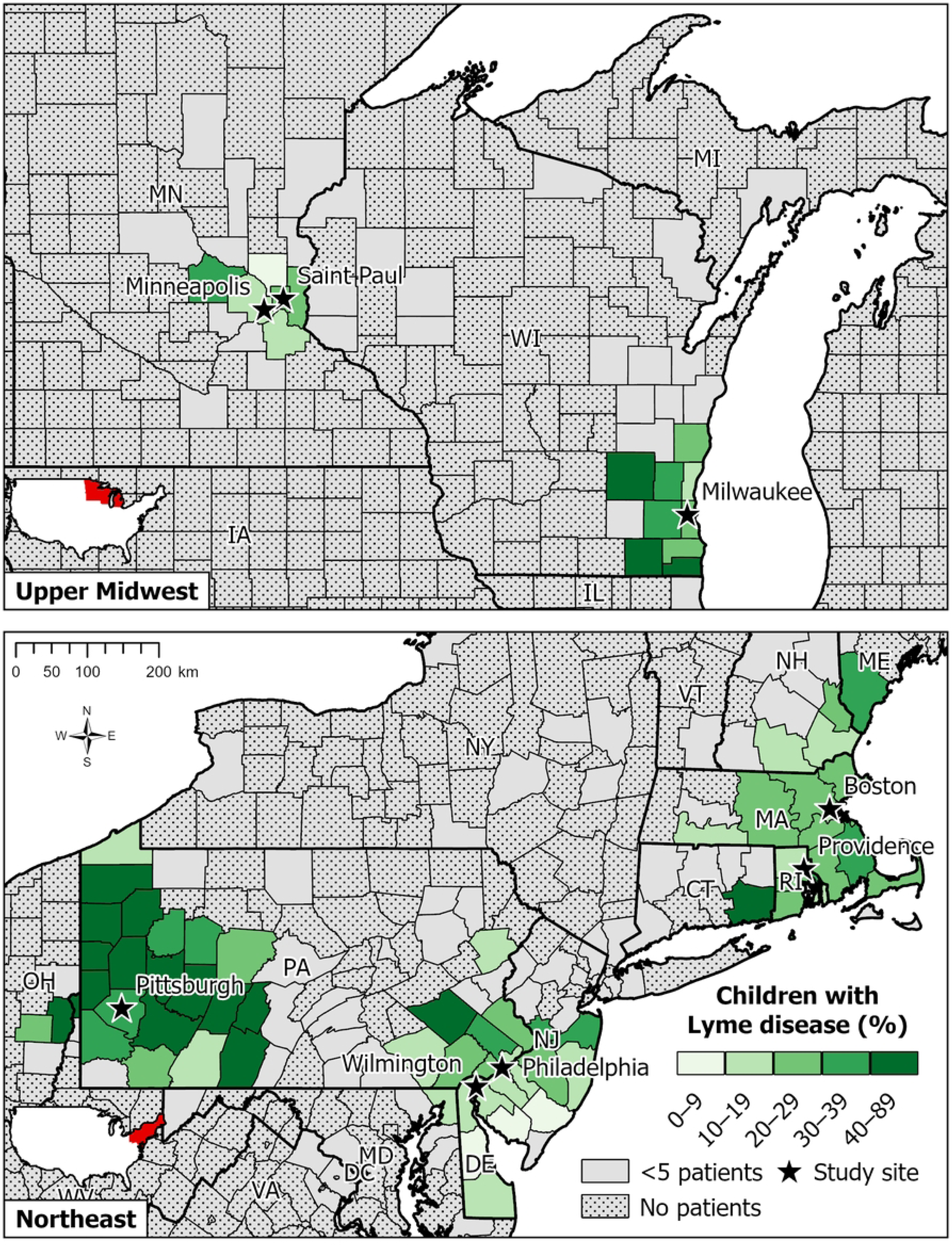
Percentage of enrolled children with Lyme disease by county of residence.

Compared to clinical mimics in our study sample, children with Lyme disease were more likely to present during the peak season (June to October) and live in counties that were rural and had higher socioeconomic advantage, “Lyme disease” Google search incidence, and wildland-urban interface (Table 1).

**Table 1.** Characteristics of patients with Lyme disease and clinical mimics, n (%) or mean (standard deviation).

| Characteristic | Lyme disease<br>n=1,396 (25.2%) | Clinical mimics<br>n=4,133 (74.8%) |
| --- | --- | --- |
| Male sex | 875 (62.7%) | 2,256 (54.6%) |
| Age in years |  |  |
| Under 5 | 234 (16.8%) | 962 (23.3%) |
| 5 to 9 | 624 (44.7%) | 1,336 (32.3%) |
| 10 to 14 | 374 (26.8%) | 1,040 (25.2%) |
| 15 and over | 164 (11.7%) | 795 (19.2%) |
| Race and ethnicity |  |  |
| Black non-Hispanic | 106 (7.7%) | 487 (11.9%) |
| Hispanic | 83 (6.0%) | 501 (12.2%) |
| Other | 80 (5.8%) | 276 (6.7%) |
| White non-Hispanic | 1,113 (80.5%) | 2,838 (69.2%) |
| Peak Lyme disease season (June to October) | 951 (68.1%) | 2,359 (57.1%) |
| Urban-rural residence |  |  |
| Urban | 1,225 (87.8%) | 3,700 (89.5%) |
| Suburban | 89 (6.4%) | 289 (7.0%) |
| Rural | 82 (5.9%) | 144 (3.5%) |
| Social Vulnerability Index (recoded high to low) | 0.72 (0.2) | 0.66 (0.2) |
| Lyme disease incidence percentile | 91.8 (4.6) | 91.3 (5.8) |
| “Lyme disease” Google search incidence | 887.1 (238.8) | 813.4 (246.4) |
| Wildland-urban interface (%) | 39.4 (17.1) | 34.7 (19.1) |

Among 5,483 patients (99.2% of study population) without missing covariates in the adjusted model, rural residence, higher socioeconomic status, higher Google search incidence, and higher wildland-urban interface were independently associated with Lyme disease after adjusting for patient demographics (Table 2). Although we included local disease incidence in the adjusted model due to its clinical relevance, it was the only predictor to show a mixed but insignificant association across models. The variance inflation factor was < 2 for all predictors, indicating no substantial multicollinearity.

**Table 2.** Unadjusted and adjusted odds ratios (OR/aOR) and 95% confidence intervals (CI) of county risk factors for Lyme disease.

| Characteristic | Unadjusted |  | Adjusted<sup>1</sup> |  |
| --- | --- | --- | --- | --- |
|  | OR | 95% CI | aOR | 95% CI |
| Urban-rural residence |  |  |  |  |
| Urban | Ref | Ref | Ref | Ref |
| Suburban | 1.0 | 0.7–1.5 | 1.0 | 0.7–1.4 |
| Rural | 2.0 | 1.3–3.0 | 1.9 | 1.3–2.9 |
| Social Vulnerability Index (recoded high to low) | 1.4 | 1.2–1.6 | 1.3 | 1.1–1.4 |
| Lyme disease incidence percentile | 1.1 | 1.0–1.2 | 1.0 | 0.9–1.0 |
| “Lyme disease” Google search incidence | 1.2 | 1.1–1.3 | 1.1 | 1.0–1.2 |
| Wildland-urban interface (%) | 1.3 | 1.1–1.5 | 1.2 | 1.0–1.4 |
| <sup>1</sup>Adjusted for sex, age, race and ethnicity, and peak Lyme disease season. |  |  |  |  |

The intraclass correlation coefficient for an empty model with no predictors was 0.11, suggesting that our observations were clustered within counties and a multi-level analysis was appropriate. The coefficient for the adjusted model decreased to 0.05, indicating that only 5% of the total variance in the odds of Lyme disease was due to unobserved county differences. Although the selected county predictors did not fully account for county variations in Lyme disease, the unaccounted variance was low, demonstrating that our results were reliable.

The results of the sensitivity analysis excluding local disease incidence and Google Trends (S1 Table) were not materially different and did not change our conclusions.

## Discussion

The increasing incidence and expanding geography of Lyme disease, apparent from decades of surveillance [31], demonstrates that geographic context is needed to understand risks. In our large multi-center cohort of children undergoing evaluation for Lyme disease, rural residence, higher socioeconomic status, more frequent “Lyme disease” Google searches, and greater intermingling of houses and wildland vegetation were associated with Lyme disease after controlling for individual characteristics. While these risk factors can inform initial decision-making, we must continue to rely on clinicians to maintain a high level of suspicion across the risk spectrum to ensure all children with Lyme disease are accurately diagnosed and treated.

Consistent with previous studies, children living in rural counties were twice as likely to have Lyme disease [11,14,15,19]. As enrolling centers were located in urban centers, we were not surprised that only a minority (< 5%) of children resided in rural counties. About half (54%) of rural children lived in Pennsylvania, a newer region of high Lyme disease endemicity [47]. The risk of Lyme disease for suburban children was no different than the urban reference group. Overall, while rural children were more at risk, Lyme disease occurred across the urban-rural gradient. While exposure is more likely in nonurban areas with more forest and outdoor recreational and occupational activities, research has shown that even highly urban areas are not immune to tick activity [48]. Lyme disease continues to increase in incidence and geographic range as *Ixodes* spp. ticks expand into new geographic areas, including suburban and urban areas where green space patterns favor tick populations and human-tick encounters [7,48–50]. Expanding urban environments that interface with wildland vegetation increase tick-borne disease risks for populations who are less aware of disease manifestation and prevention [7,51].

Greater socioeconomic advantage was a risk factor for Lyme disease, contrasting with classic income-health relationships for most diseases [52] but agreeing with other studies of Lyme disease [10–13]. More affluent families may live in lower density areas with woodland interface, where ticks are more common [7,53,54], and have greater access to outdoor recreational activities such as outdoor sports, hiking, and camping. This finding should not be interpreted as low socioeconomic status having a protective effect against Lyme disease. Another study found that patients with lower socioeconomic status and less health care access had a higher risk of disseminated Lyme disease, possibly due to delays in care-seeking, diagnosis, and treatment [55]. Despite the observed association, clinicians should continue to evaluate children across the socioeconomic spectrum for suspected Lyme disease.

Greater proximity of human habitation to wildland vegetation was associated with increased risk of Lyme disease, illustrating the important role of the built environment in shaping health. Lyme disease risk is high in fragmented forest ecosystems, which are a consequence of suburban sprawl [56]. Forest fragmentation metrics on their own have shown mixed associations with Lyme disease [10,48,57–59]. Limited studies have explored the relationship between wildland-urban interface and Lyme disease [18,59,60]. Our finding suggests that this measure captures the intersection of tick habitat and human activity.

Surprisingly, local Lyme disease incidence, as reported by CDC, was not associated with Lyme disease in our prospective pediatric cohort. The average incidence (expressed as a percentile rank) was > 90% among cases and clinical mimics, indicating that in very high-risk areas, population-level incidence may not be enough to risk-stratify individual children with suspected Lyme disease.

Google search incidence for “Lyme disease” was associated with increased risk of disease. Internet search data are now widely used to capture public interest and concern and explore spatiotemporal patterns in health information-seeking behavior. Prior studies have shown that temporal peaks in tick-borne disease-related searches precede those for reported cases, and that spatial patterns in searches approximate the geographic incidence and spread of disease over time [20,21,61–63]. As disease risks change with climate change and other human activities [64], Google search data may provide health care professionals with early warnings of disease activity and diagnostic decision support in their geographic service areas. An advantage of the tool is that search trends may be queried for any time range (e.g., past day, week, or month), providing a more real-time signal not available from other data sources.

Our study has important limitations. First, area-level measures of social determinants of health, while easily accessible and often used as proxies in public health research, may fail to capture individual-level risks [65]. Where someone lives influences, but does not fully determine, the complex factors impacting individual knowledge, attitudes, and practices regarding Lyme disease [66]. Area-level measures should be incorporated carefully to ensure clinical assessments are not based on geographic location alone. Second, we used county as our geographic unit of exposure as most risk factors were not measured at the ZIP Code level. Socio-ecological risks such as degree of urbanization and tick habitat proximity may vary substantially within counties. In addition, a patient’s county of residence may differ from the county of tick exposure due to multiple residences, travel, and occupational or recreational exposure. Third, while Google Trends provides a promising tool for public health surveillance, we were unable to distinguish the clinical significance of “Lyme disease” searches, which may reflect diagnosis-seeking from affected families, general curiosity, or recent media attention. The lack of county-level search trends and extrapolation of market area trends to counties was also a limitation. Although not routinely available, more detailed internet search data may provide more accurate forecasting [20] and should be explored further. Fourth, the population-level Lyme disease incidence variable was limited by incomplete data and reporting changes over a ten-year period, and our approach to addressing these limitations was imperfect. Despite the lack of statistical significance, local epidemiological risk is still valuable for diagnostic decision-making and may be a more obvious metric for clinicians with no other knowledge of geographic risk factors of Lyme disease. The proliferation of public health dashboards like the Lyme and Tickborne Diseases Dashboard [31,32] makes it easier than ever for clinicians to find and visualize disease data. Lastly, we enrolled children evaluated in emergency departments located in Lyme disease endemic areas. Therefore, our findings may not be generalizable to children in outpatient clinics or in lower-risk or emerging areas. The locations of these emergency departments produced a mostly urban cohort, with only 7% and 4% of patients residing in suburban and rural counties, which limited our ability to explore urban-rural differences. For reference, in Lyme disease endemic states, 25% and 12% of the population live in suburban and rural counties [27,30]. Future study is needed to evaluate risk factors in diverse practice settings.

## Conclusion

In this large multi-center cohort of children with suspected Lyme disease, we identified important socio-geographic risk factors for Lyme disease. Clinicians may leverage these factors to inform clinical decision-making for children with suspected Lyme disease. Future study incorporating geographic risks alongside individual clinical factors may further augment risk assessment for clinical practice.

## Supporting information

**S1 Table. Sensitivity analysis: unadjusted and adjusted odds ratios (OR/aOR) and 95% confidence intervals (CI) of county risk factors for Lyme disease.**

## Data Availability

Due to ethical and privacy reasons, patient data cannot be made publicly available for reproducibility as the analysis relies on personally identifiable date and location information. Please contact Lise E. Nigrovic for data sharing requests. Area-level datasets are publicly available from the National Center for Health Statistics (https://www.cdc.gov/nchs/data-analysis-tools/urban-rural.html), U.S. Centers for Disease Control and Prevention (https://www.atsdr.cdc.gov/place-health/php/svi/svi-data-documentation-download.html, https://www.cdc.gov/lyme/data-research/facts-stats/lyme-disease-case-map.html), and the Silvis Lab at the University of Wisconsin-Madison (https://silvis.forest.wisc.edu/data/wui-change). Researchers may request Google Trends API access at https://support.google.com/trends/contact/trends_api or obtain comparable data from the public Google Trends website (https://trends.google.com/trends).

